# Covid-19 vaccination coverage and break through infections in urban slums of Bengaluru, India: A cross sectional study

**DOI:** 10.1101/2021.11.21.21262716

**Authors:** Sunil Kumar D.R, Srividya J, Apoorva E Patel, Vidya R

**Affiliations:** Akash Institute of Medical Sciences and Research Centre, Bengaluru, India

**Keywords:** COVID-19 vaccination, Break through infections, Vaccine hesitancy, Adverse events COVID vaccination, Urban slums

## Abstract

**Background:** The ongoing pandemic of Corona virus disease 2019(covid-19) is caused by severe acute respiratory syndrome Corona virus 2(SAR-COV-2). The world health organization declared it as public health emergency of international concern on January 2020, and later declared as pandemic on 11 March 2020.One of the high-risk groups for COVID-19 disease are people residing in urban overcrowded slums and as most of the population is migrant, they are less aware of the pandemic and have less access to health care facilities.

Vaccinating these high-risk groups can decrease disease burden and control the ongoing pandemic.

**Objectives:** 1] To estimate COVID 19 vaccination coverage 2] To assess the factors responsible for COVID - 19 vaccination coverage and vaccine hesitancy 3] To study AEFI pattern following COVID-19 vaccination 4] To determine the prevalence of breakthrough infections after COVID - 19 Vaccination in urban slums of Bengaluru, India.

**Methodology:** A community based cross sectional study was conducted in Urban slums belonging to Urban Health and Training Centre, Department of community medicine, Akash Institute of Medical Sciences and Research Centre, Bengaluru Rural District, Karnataka, India. After obtaining Institutional ethical clearance and informed consent from study participants, data was collected from 1638 participants, fulfilling inclusion criteria using a predesigned, pretested, structured questionnaire. Data was entered in Microsoft excel and analyzed using SPSS version 24. Chi square test and Fischer’s exact test was applied and p <0.05 considered as statistically significant.

**Results:** In the present study, 35.5% (583 out of 1638) of the study participants had taken COVID Vaccine, of which 533 (91.42%) were partially vaccinated and remaining 50 (8.5%) were fully Vaccinated. Majority i.e., 98.45% have taken vaccine at Govt health centers. 63.65% vaccinated with Covishield reported adverse events, whereas 18.6% vaccinated with Covaxin reported adverse events. Adverse events were more likely to be reported by women (74.7%) compared to men (58.6%), this observation was consistent across all age groups. Vaccination coverage was high among 18 – 45 years age group (37.75%), males (64.86%), Christians (47.05%) followed by Hindus (43.56%), graduates (95.67%), clerical and skilled workers (70.75%), Upper middle socioeconomic class (72.41%). This difference was statistically significant. Our study reported Break through infections in 7 out of total 583 vaccinated with a prevalence of 1.2%. The break through infections was very high among partially vaccinated (85.71%) as compared to fully vaccinated individuals (14.28%). This was observed among those vaccinated with Covaxin only.

**Conclusion:** The COVID vaccine coverage was low in urban slums. The prevalence of Break through infections in our study was higher as compared to available data/reports in the country. Break through infections was very high among partially vaccinated as compared to fully vaccinated individuals. This study on break through infections on COVID vaccination is first study in South India on general population. The most important factor for vaccine hesitancy is the occurrence of mild or serious adverse effects following immunization, and this may be the biggest challenge in the global response against the pandemic.

## INTRODUCTION

The ongoing pandemic of Corona virus disease 2019(covid-19) is caused by severe acute respiratory syndrome Corona virus 2(SAR-COV-2). The virus was first identified in December 2019 in Wuhan, China. The world health organization declared it as public health emergency of international concern on January 2020, and later declared as pandemic on 11 March 2020. As of 9 July 2021, more than 185 million cases have been confirmed, with more than 4.01 million confirmed deaths attributed to covid-19 making it one of the deadliest pandemic in the history.^[1]^

In India, 3crore confirmed cases are reported, out of which presently 4.5lakh cases are active with 405939 deaths. In Karnataka state, there are 28lakh confirmed cases with 37000 active cases and 35731 deaths. India began its vaccination program on 16 January 2021, by operating through 3006 vaccination centres across the country. As on 9 July 2021, 372196268 doses of vaccine are given across the country among which 2.5crore vaccine are administered in Karnataka state.^[2]^

Immunization is the process where in a person is made immune or resistant to an infectious disease, typically by administering vaccine and stimulating human body to the pathogen there by making immune system competent enough to combat the infection. From history we can easily make out how vaccines helped in eradicating small pox, eliminating polio from India. Vaccines also help in reducing disease burden, mortality and morbidity associated with a disease. By vaccinating two-third of the population we can reach HERD IMMUNITY THRESHOLD thereby preventing further spread of the disease and also break the transmission chain.^[3]^

With the COVID vaccination drive widening across India, a ‘class divide seems to be opening up, almost every centre in big cities is reporting more recipients from wealthier quarters. The reasons for this divide could range from poor access to smartphones, digital illiteracy, high priced vaccines to transport issues and vaccine scepticism. One of the high-risk groups for COVID-19 disease are people residing in urban overcrowded slums as most of the population is migrant, they are less aware about the pandemic and have less access to health care facilities. Approximately 1 billion people live in such settlements globally, more than half of this population is present in Asia and almost 1/5^th^ in India. ^[4]^ Vaccinating these high-risk groups can decrease disease burden and control the ongoing pandemic.

Vaccine hesitancy has been a long-standing issue amongst slum dwellers and earlier vaccine drives for polio, TB, influenza have always faced tough situations when it came to urban poor communities who mistrust the system. The COVID vaccination drive has until now, been over reliant on digital registration. A large quota of vaccinations being provided by private clinics and hospitals led to a significant class divide in the earlier days of the vaccine program. Issues like co-morbidities and age were being given preference but the factors of livelihood, vulnerable and exposed livelihoods were completely omitted. Though the vaccination drive is widening across the county, Vaccination centres are fairly unknown if one does not log onto the CoWin website and locate the centre. This is a major hindrance for the urban poor communities.

Till now COVID – 19 Vaccination coverage has not been widely addressed in the Indian urban slums context. Addressing this aspect of vaccination will help the policymakers to undertake appropriate measures to improve vaccine acceptance, coverage and reach desired national targets and to combat the COVID 19 pandemic.

## OBJECTIVES

- 1] To determine the COVID 19 vaccination coverage in urban slums of Bengaluru, India.
- 2] To assess the factors responsible for COVID 19 vaccination coverage and vaccine hesitancy in urban slums of Bengaluru, India.
- 3] To study AEFI pattern following COVID-19 vaccination in urban slums of Bengaluru, India.
- 4] To determine the prevalence of breakthrough infection after COVID 19 Vaccination in urban slums of Bengaluru, India.

## METHODOLOGY

**Study design**- Community based cross sectional study.

**Study period**- Four months between June to Sept 2021.

**Study area**- Urban slums belonging to the field practice area of Urban Health and Training Centre (UHTC), Department of Community Medicine, Akash Institute of Medical Sciences and Research Centre, Devanahalli, Bengaluru, Karnataka, India.

### Study population

Inclusion criteria:

Adults aged 18 years and above, residing for more than 1 year in the above-mentioned urban slums and willing to participate in the study.

Exclusion criteria:

1. Individuals aged < 18 years.
2. Pregnant and lactating women.

### Sample size

Based on Indian census 2011, family folders maintained at UHTC, Anganwadi, UHTC consists of 11 slums with total 1171 households with a population of 6029, of which 3468 are >18yrs, and 987 are pregnant and lactating women. So the population above 18 years excluding pregnant and lactating women was around 2481.Considering the migrant nature of the slum population, current family folder records maintained during the study period at Anganwadi and UHTC was considered for calculating the sample size. All the adults above 18 years residing in the 11 slums for more than 1 year (excluding pregnant and lactating women) were considered, and the final sample size was 1638.

Institutional ethical clearance was obtained from Institutional Ethical Committee. Data was collected from 1638 participants, fulfilling inclusion criteria. After taking informed consent from study participants, data was collected using a predesigned, pretested, structured questionnaire. Questionnaire consisted of details on socio demography, vaccination, adverse events following covid vaccination and other epidemiological details. The study participants were assured of confidentiality / anonymousity.

### Statistical analysis

Data was entered in Microsoft excel and analyzed using SPSS version 24. Appropriate test of significance was applied to determine any statistical association [chi square test and Fischer’s exact test] between dependent and independent variables. [p<0.05 considered as statistically significant].

## RESULTS

The present study was conducted among 1638 study participants. The socio demographic details of the study participants given in Table 1.

**TABLE1:**
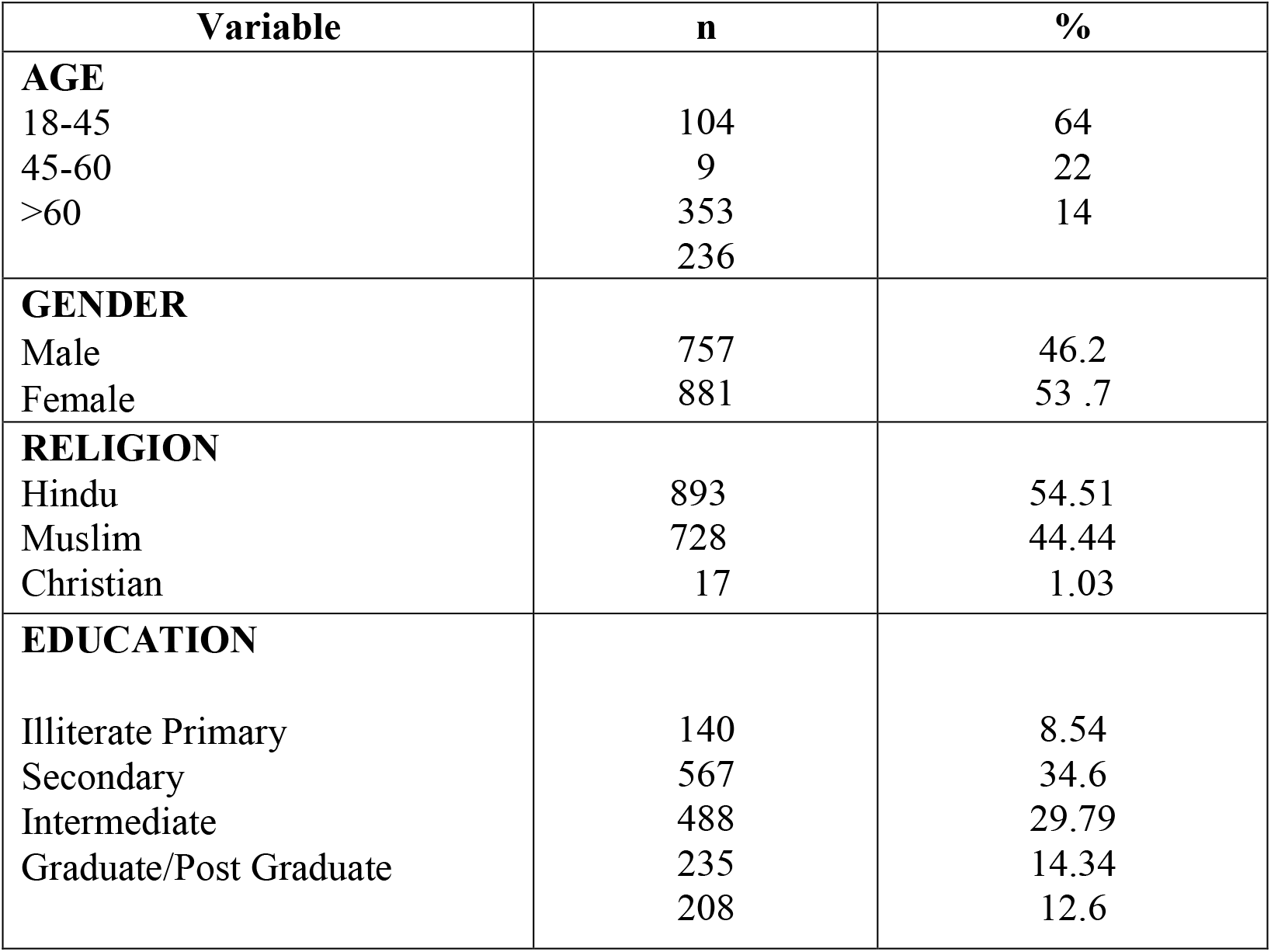
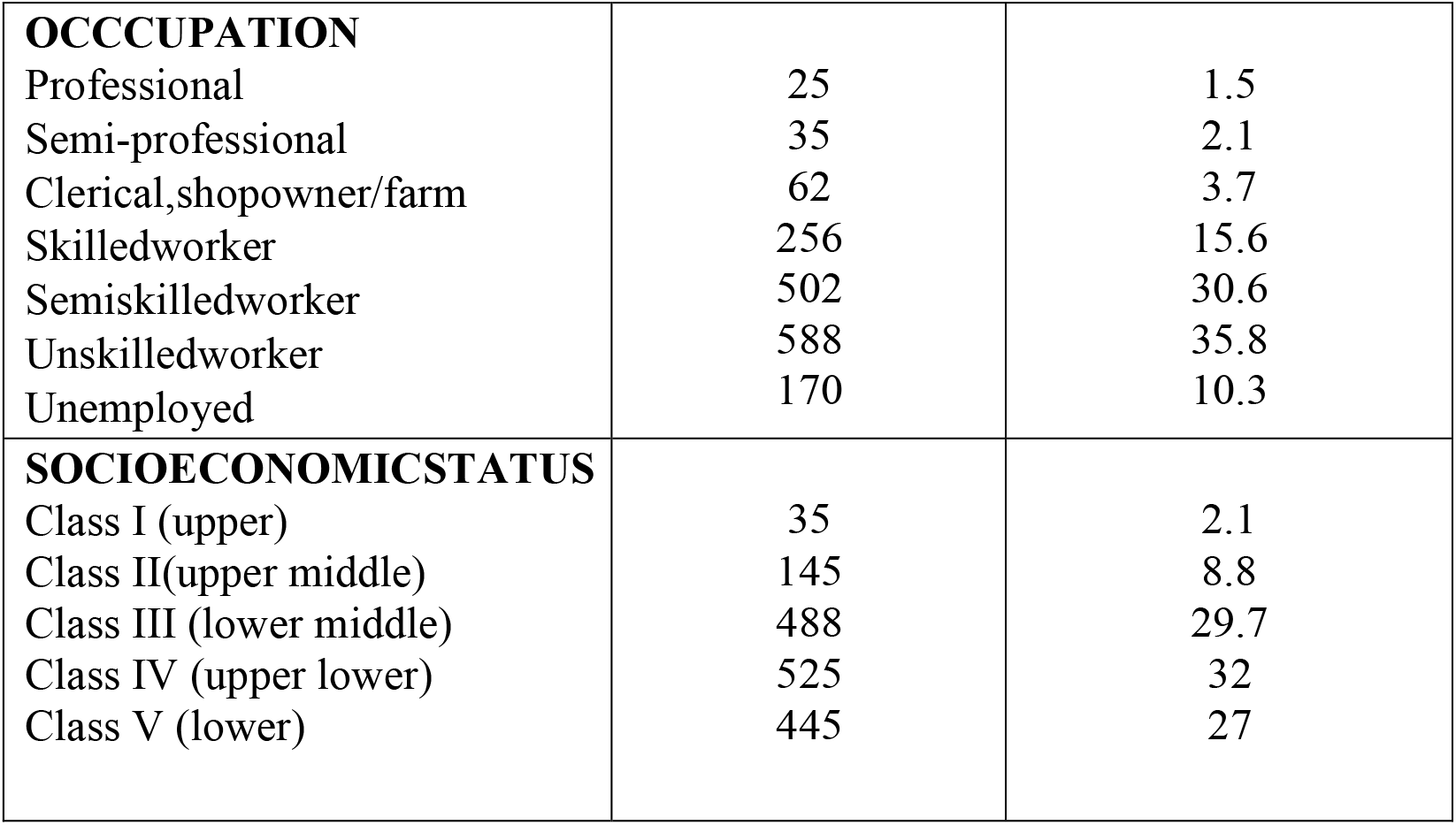
SOCIO DEMOGRAPHIC DETAILS:

35.5% (583 out of 1638) of the study participants had taken COVID vaccine. Partially vaccinated - 533 (91.42%), Fully Vaccinated - 50 (8.5%) [Fig 1]. Majority i.e., 98.45% have taken vaccine at Govt health centres.

**FIGURE 1:**
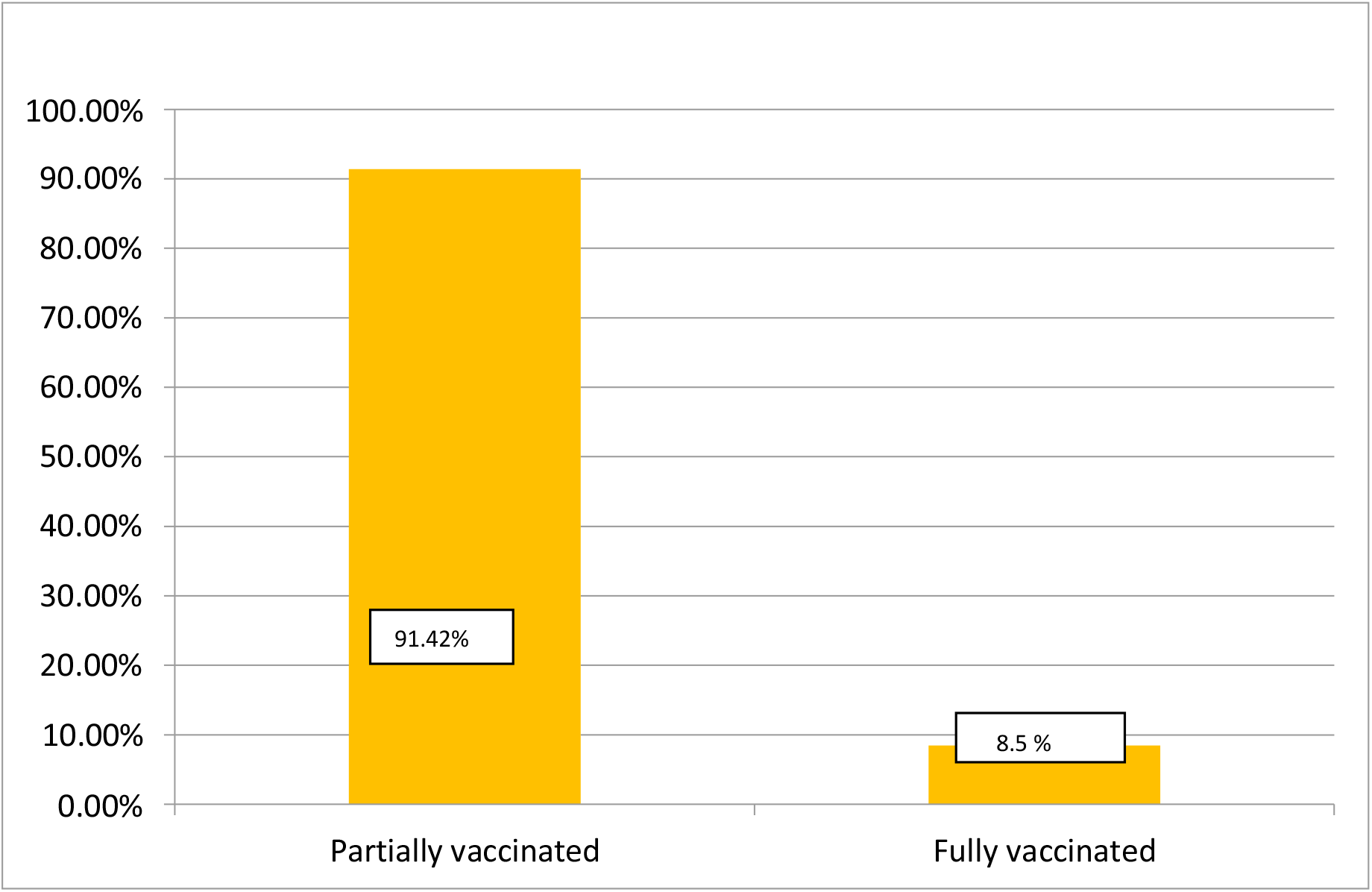
COVID-19 VACCINATION COVERAGE.

**FIGURE 2:**
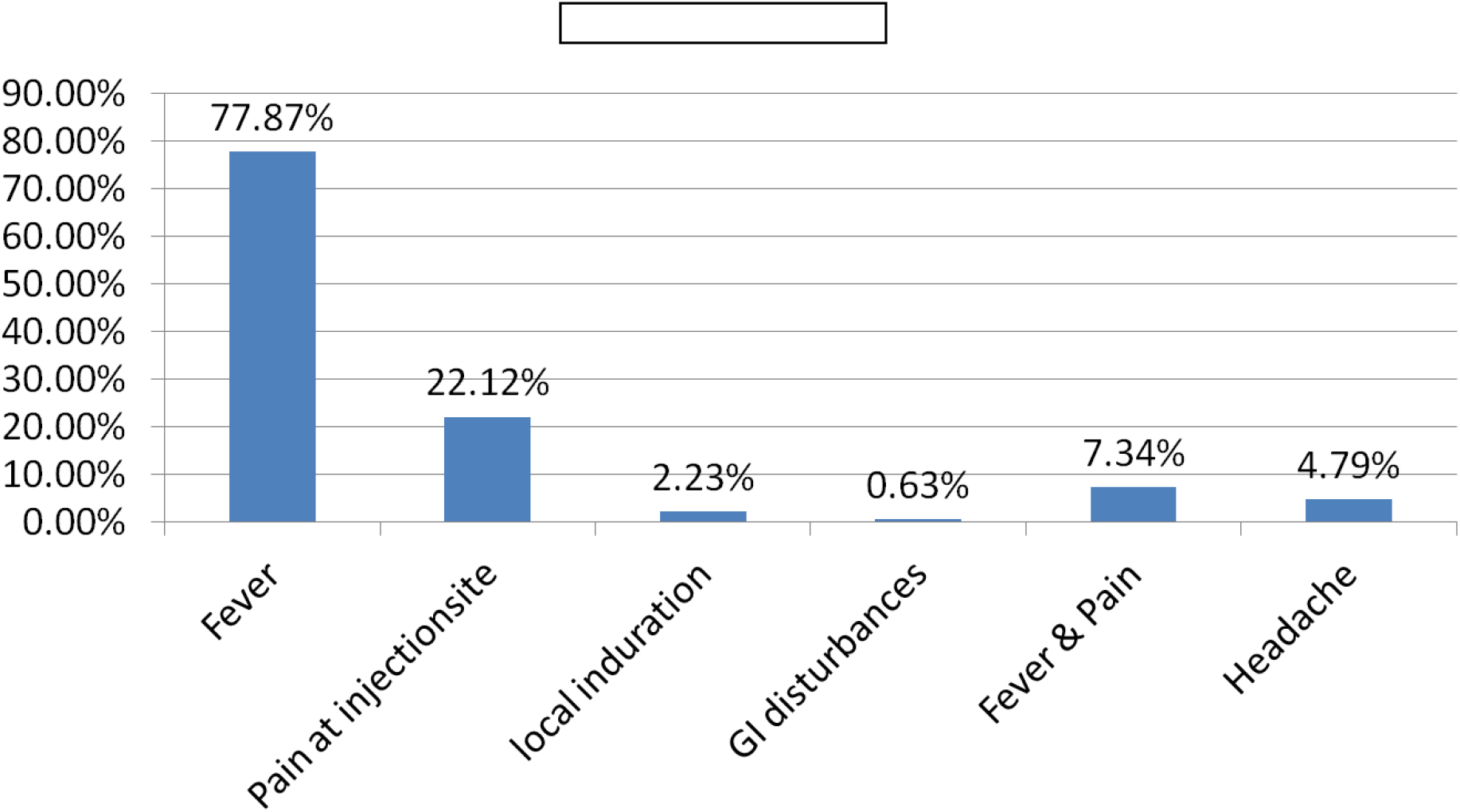
ADVERSE EVENTS FOLLOWING COVID-19 VACCINATION:

Reasons reported for unwillingness to vaccinate are-Fear of Side-effects (36%), doubts on vaccine safety (20%), non-availability of vaccine (18%), fear of infertility (12%), assumption of not being at risk of getting COVID infection (14%).

In the present study, vaccination coverage was high among 18 – 45 years age group (37.75%), males (64.86%), Christians (47.05%) followed by Hindus (43.56%), graduates (95.67%), clerical and skilled workers (70.75%), Upper middle socioeconomic class (72.41%). This difference was statistically significant. [Table 2]

**TABLE 2:**
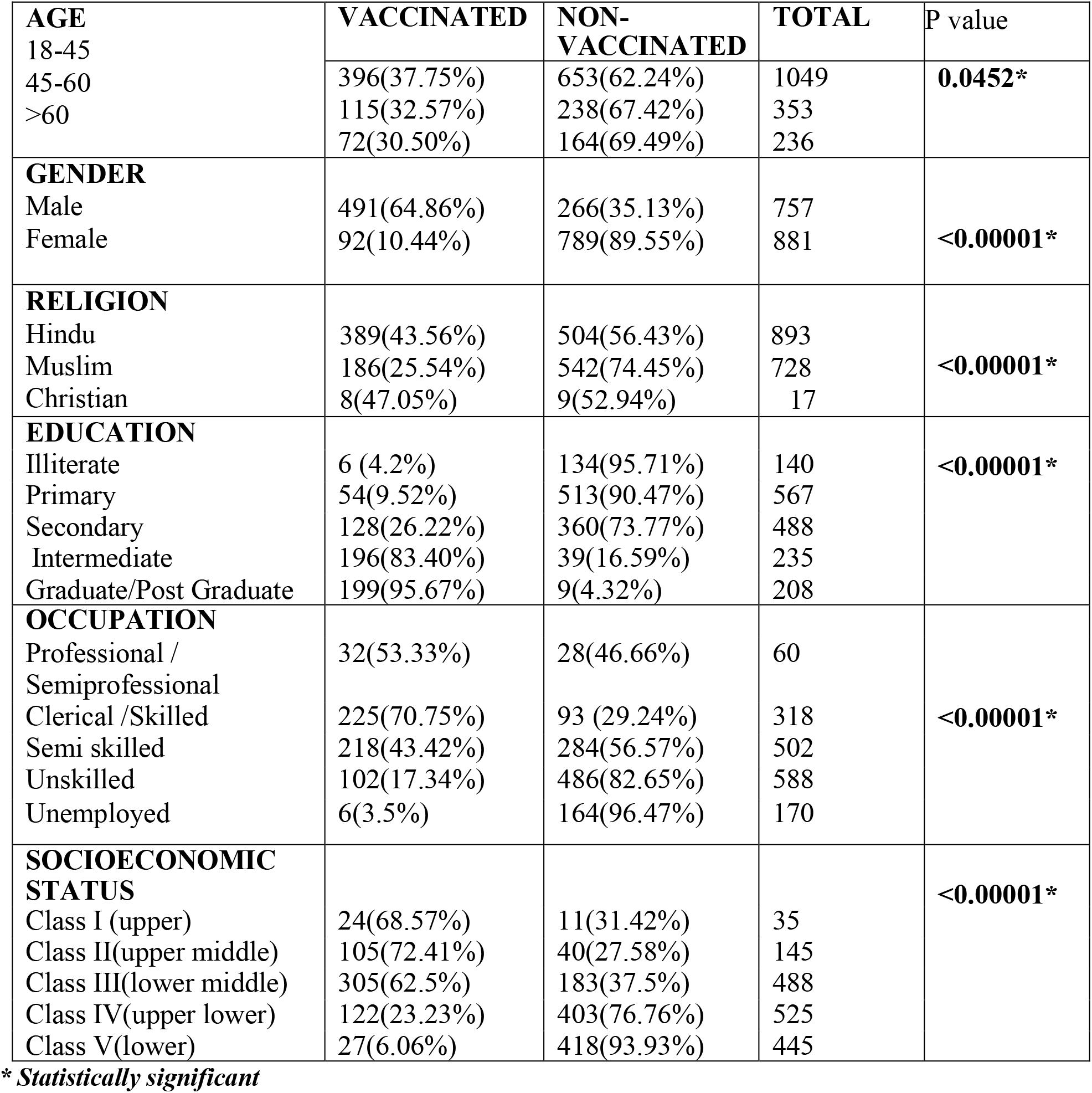
FACTORS ASSOCIATED WITH VACCINATION COVERAGE AMONG VACCINATED AND NON-VACCINATED:

In the present study, majority of the study participants above 45 years and majority of the females have taken Covishield. Study participants belonging to Upper and Upper middle class have taken Covaxin and study participants belonging to lower middle and lower socioeconomic class have taken Covishield. This difference was statistically significant.

AEFI was reported by 53. 68% (313 out of 583) vaccinated study participants. 289 out of 454 (63.65%) study participants vaccinated with Covishield reported AEFI, whereas 24 out of 129 (18.6%) study participants vaccinated with Covaxin reported AEFI.

AEFI were more likely to be reported by women (74.7%) compared to men (58.6%), this observation was consistent across all age groups. Women were more likely to report AEFI severe enough to prevent working for a day (27% vs. 15% in males) and the need to take pain relievers (70% vs. 51% in males). Among those who reported AEFI, 79% noticed them within the first 12 hours after vaccination. Women had slightly longer duration of AEFI for more than 2 days compared to males (less than 1 day).

Older people had later onset of AEFI, occurring at an average of 13.4 hours (70-79 years), compared to 10 hours in younger age groups (20-29 years) following vaccination. The duration of AEFI decreased with advancing age, ranging from an average of 1 ½ day in younger age groups (20-29 years) to less than 1 day in older age groups (70-79 years) In the present study, more adverse events were reported with Covishield compared to Covaxin. This difference was statistically significant. [Table 3]

**TABLE 3.** ADVERSE EVENTS REPORTED WITH RESPECT TO THE TYPE OF VACCINE TAKEN:

| VACCINE | ADVERSE EVENTS REPORTED | ADVERSE EVENTS NOT REPORTED | TOTAL | P <0.00001* |
| --- | --- | --- | --- | --- |
| COVISHIELD | 289 (63.65%) | 165 (36.34%) | 454 |  |
| COVAXIN | 24(18.60%) | 105(81.39%) | 129 |  |
| TOTAL | 313 | 270 | 583 |  |

\* statistically significant

In the present study, break through infection was reported among 7 out of 583 study participants who got Covid vaccine with a prevalence of 1.2%. The break through infection was reported from those vaccinated with Covaxin. This was statistically significant [Table 4]. Majority (85.71%) of the break through infections were reported among partially vaccinated study participants and 14.28% of the breakthrough infections reported among fully vaccinated study participants. However, this difference was not statistically significant. [Table 5]

**TABLE 4:** BREAKTHROUGH INFECTIONS REPORTED AND TYPE OF VACCINE.

| VACCINE | BREAKTHROUGH INFECTION REPORTED | NOT REPORTED | TOTAL | p<0.05* |
| --- | --- | --- | --- | --- |
| COVISHIELD | 0 | 454 (100%) | 454 |  |
| COVAXIN | 7 (5.4%) | 122(94.57%) | 129 |  |
| TOTAL | 7 | 576 | 583 |  |

\* *statistically significant*

**TABLE 5:** BREAKTHROUGH INFECTIONS REPORTED AND VACCINATION STATUS.

| Break Through Infections | Partially Vaccinated | Fully Vaccinated | TOTAL | P value 0.58 |
| --- | --- | --- | --- | --- |
| REPORTED | 6(85.71%) | 1(14.28%) | 7 |  |
| NOT REPORTED | 527(91.49%) | 49(8.5%) | 576 |  |
| TOTAL | 533 | 50 | 583 |  |

## DISCUSSION

In the present study, 35.5% (583 out of 1638) of the study participants had taken COVID Vaccine of which 533(91.42%) were partially vaccinated and remaining 50(8.5%) were fully vaccinated. This correlates with the state level and national level data during the study period.^[5]^

In a study done by Lazarus et al in 19 countries, Asian countries showed more than 80% acceptance of the COVID 19 vaccine highest being in China (90%) and the lowest acceptance was found to be in Russia (55%).^[6]^ Sallam et al had reported that in Arab countries, COVID 19 vaccine acceptance was 29.4% and most important reason for refusal (23.4%) was the conspiracy theories that it could lead to infertility. ^[7]^ In studies conducted in the US and Italy acceptance was between 57.6% - 68.6% and reason for vaccine hesitancy was doubts about the effectiveness and potential side effects.^[8-10]^

In the present study reasons reported for unwillingness to vaccinate are - fear of side effects (36%), doubts on vaccine safety (20%), non-availability of vaccine (18%), fear of infertility(12%), assumption of not being at risk of getting COVID infection(14%). The study participants being Socio-economically disadvantaged group and having inadequate knowledge regarding COVID-19, inadequate preventive measures against the virus infection ^[11]^., could have possibly contributed to their negative attitude toward unwillingness to vaccine. Fear of losing a day’s pay to get the jab, or possible adverse events that could force them to skip work for longer, lack of knowledge about - where to get vaccinated, how to register for the jab, documents to be furnished, other eligibility criteria to the large populations of slum dwellers in cities. Financial distress is also a major factor hindering accessibility to the vaccine or general health services. The fear of losing even a day’s salary is very real for many people who have to pay back huge debts accumulated. There is a greater fear about the vaccines amongst women who think pregnancy and fertility gets affected, and are therefore less willing to be administered.

Close results have been found in a similar survey in Mumbai slum in India, which also demonstrated a 20% unacceptance of vaccine for COVID-19 among the slum dwellers ^[12] [13]^ Another citizen-survey platform in Delhi found that about 69% of respondents saw no urgent need to get immunized, key reasons for hesitancy included limited information about side-effects, efficacy levels, and perceived high immunity levels.^[14]^ This resonates with a global survey (in collaboration with the World Economic Forum) conducted in October 2020 of more than 18,000 adults from 15 countries that reported confidence being down by 4 points compared to the previous round in August 2020.^[15]^ Vaccination intent declined in 10 of the 15 countries, most of all in China, Australia, Spain, and Brazil. Globally, 10% reported that they were against vaccines in general, including 14% in India and South Africa.

Out of those who got vaccinated, 98.45% walked to government vaccine facilities and got registered. Only 1.5% of them registered themselves on the CoWin website and the Aarogya Setu application. The issue of the digital divide is very clearly visible in this piece of data.

In the present study, vaccination coverage was high among 18 – 45 years age group (37.75%), males (64.86%), Christians (47.05%) followed by Hindus (43.56%), graduates (95.67%), clerical and skilled workers (70.75%), Upper middle socioeconomic class (72.41%). This difference was statistically significant. The rate of unwillingness to take a COVID-19 vaccine was almost double among the 60+ years population compared to other age groups. The percentage of vaccine hesitancy was also highest among the older age group. The aging population of this study held a negative attitude towards vaccination for COVID-19, which is concerning as these groups are the most vulnerable to the adverse outcome of coronavirus infection. The older population is also substantially lagged in literacy rate than other age-groups, which can mold their perception and knowledge regarding COVID-19 and thus influence the decision for vaccination. This differs from the vaccine acceptance rate among the older population of the US and Saudi-Arabia, who found a higher prevalence of acceptance in this group. ^[17, 18]^ Furthermore, the respondents’ education and income increased the percentage of COVID19 vaccine acceptance with growing years of schooling and family income. This is consistent with the findings from a global vaccine acceptance survey involving 19 countries where respondents with high income and higher education were more likely to vaccinate against COVID-19. ^[19]^

In the present study majority (77.87%) reported fever followed by pain at the injection site (22.12%) as the adverse events after vaccination. This is contrary to findings of other studies which reported pain at the injection site as the major adverse event. There was a clear linear correlation between age and side effects, suggesting that vaccine reactogenicity declined with age. In the youngest age group (20-29 years) 81.3% developed side effects, while only 7.4% of those over 80 years reported any side effects. Vaccine reactogenicity is known to correlate with transient elevation of inflammatory cytokines, but is not considered a reliable sign of a desirable immune response. ^[20]^ Women were more likely to develop adverse events. The onset of adverse events was slightly earlier and the duration slightly longer in this group. This observation was consistent across all age groups. One of the reasons why women tend to experience more adverse events could be because of the way hormones interact with the immune system make up. Heightened estrogen levels may lead to more inflammatory reactions and increase the duration of adverse events as well. The findings of the study correlated with results from published trials of vaccines. In the phase 2/3 trial of Astra-Oxford ChAdOx1 nCoV-19, at least one systemic symptom was reported following vaccination with the standard dose by 86% participants in the 18–55 years group, 77% in the 56–69 years group, and 65% in the 70 years and older group. ^[21]^ While discussing post vaccination experience, it is noteworthy that placebo injections produce comparable symptoms. In the phase 3 trial of Pfizer-Biontech vaccine, the incidence of headache following vaccination was 42% in the vaccine group and 34% in those who received saline placebo. ^[22]^ This has been termed the nocebo effect, which results from enhanced anticipation of negative outcomes from an intervention. ^[23]^

This study did not measure post vaccination antibody response. Hence it is not possible to infer whether the muted side effects among older people was a sign of immune senescence. Although side effects are known to correlate with neutralising antibody levels during COVID-19 ^[24]^, the presence of side effects following vaccination does not reliably predict antibody response^[25]^. The frequency of using paracetamol to reduce side effects decreased from 71% in the 20-29 age group to 16% in the 80-90 age group. This correlated with the side effects frequency in these subgroups. Although the use of paracetamol to alleviate postvaccination discomfort is considered acceptable, routine prophylactic use of pain relievers is not recommended as there is evidence of blunted immune response as a result. ^[26,27]^

In the present study, 63.65% study participants vaccinated with Covishield reported adverse events, whereas only 18.6% study participants vaccinated with Covaxin reported adverse events. The difference was statistically significant. High percentange of adverse events reported with Covishield may be be due to the fact that it is a viral vector vaccine that uses an Adenovirus found in Chimpanzees,ChADox1, to deliver spike proteins and mounts a higher antibody response(immune response).

Our study reported Break through infections in 7 out of total 583 vaccinated with a prevalence of 1.2%. The prevalence of Break through infections in our study was higher as compared to available data/reports in the country. As per government data, during study period, 23,940 people got infected by COVID-19 after taking Bharat Biotech’s Covaxin in India, which is 0.13 per cent of the total vaccine doses delivered. Of these, 18,427 were infected after the first dose, while 5,513 were infected after the second dose. For covishield, produced by the Serum Institute of India, 1,19,172 breakthrough infections have been reported, which is 0.07 per cent of the administered doses. Of these, 84,198 got infected after the first dose, while 34,874 got infected after the second dose.^[28]^

In our study, it was observed that, break through infections was very high among partially vaccinated (85.71%) as compared to fully vaccinated individuals (14.28%). However, this difference was not statistically significant. Break through infections was observed among those vaccinated with Covaxin only in our study. As per Government data, there is not much difference in breakthrough COVID 19 infections amongst those vaccinated with Covishield and Covaxin vaccines.

### Limitations

The study has few limitations. Although this study comprehensively explored the socio demographic determinants of vaccine Coverage, the influence of essential factors like misinformation on vaccine safety and effectiveness on the intention to vaccinate was not explored in this study. Relationship with trust in the various sources of information such as healthcare sectors and media with vaccine acceptance has also not been addressed, which could also increase the study`s strength. There could be recall bias from the study participants while reporting the type and duration of adverse events following COVID vaccination.

## CONCLUSION

The COVID vaccine coverage was low in urban slums. The prevalence of Break through infections in our study was higher as compared to available data in the country. Break through infections was very high among partially vaccinated as compared to fully vaccinated individuals. This study on break through infections on COVID vaccination is first study in South India on general population. The most important factor for vaccine hesitancy is the occurrence of mild or serious adverse effects following immunization, and this may be the biggest challenge in the global response against the pandemic.

The rapid development of COVID-19 vaccine might have contributed to the emergence of concerns among the general population. Vaccine coverage may be increased once additional information about vaccine safety and efficacy is available in the public domain, preferably by a trusted, centralized source of information. In addition, all efforts must be made to curb the spread of misinformation about the vaccine. Interventional educational campaigns especially targeting the populations at a higher risk of vaccine hesitancy are therefore essential to avoid low inoculation rates.

## RECOMMENDATIONS

Rumors and vaccine hesitancy perceptions can act synergistically to shape demand, vaccine confidence, and vaccine hesitancy. In view of the fact that it is a new disease against which several candidate vaccines are being developed and licensed in a fast-tracked manner communication strategy, communication messages shall need to address four key themes: (i) product development, (ii) prioritization strategies, (iii) program rollout activities, and (iv) adverse events following immunization and adverse events of special interest.

The role of health care workers is crucial in sustaining the success of vaccination programmes. It is necessary to improve their knowledge about vaccination and stimulate them to promote vaccination practices. Capacity strengthening of health care workers should be done. Effective implementation of national surveillance programme of adverse events following immunisation is of prime importance for building evidence about vaccine safety and assuring the public that continuous monitoring is in place to help assessing any suspicion of safety issue. Mobile-based vaccine reminders can be widely used to address delays.

Although recently the central government has made walk in registration facility mandatory for all states, it is upto all local governments, CSOs and other ground staff to implement these measures as the digital divide will result in more inequality.

Additional studies to identify the barriers to covid vaccine coverage, break through infections and the populations at a higher risk for vaccine hesitancy are also critical. They will help the public health policy makers to formulate more definitive, efficient strategies that can help to implement the COVID-19 vaccination program successfully in India.

### Measures for enhancing vaccination amongst urban poor

1. Establishing Community Help desks.
2. Development of Community Volunteers.
3. Multi-lingual COWIN website.
4. Mobile vaccinations.
5. Monitoring marginalised urban poor population.

### Measures For vaccine awareness, better coordination and support to volunteers

1. Use of socio-religious mobilisers
2. Walk in Centres near slums
3. Training of Community Task Force
4. Strengthening IEC
5. Involvement of NGO/CSO volunteers:

## Data Availability

Data Availability Later

## Ethical approval

The study was approved by the Institutional Ethics Committee.

